# Thoracic Aortic Shape: A Data-Driven Scale Space Approach

**DOI:** 10.1101/2024.08.30.24312310

**Authors:** Joseph A. Pugar, Junsung Kim, Kameel Khabaz, Karen Yuan, Luka Pocivavsek

## Abstract

The scale and resolution of anatomical features extracted from medical CT images are crucial for advancing clinical decision-making tools. While traditional metrics, such as maximum aortic diameter, have long been the standard for classifying aortic diseases, these one-dimensional measures often fall short in capturing the rich geometrical nuances available in progressively advancing imaging modalities. Recent advancements in computational methods and imaging have introduced more sophisticated geometric signatures, in particular scale-invariant measures of aortic shape. Among these, the normalized fluctuation in total integrated Gaussian curvature 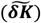 over a surface mesh model of the aorta has emerged as a particularly promising metric. However, there exists a critical tradeoff between noise reduction and shape signal preservation within the scale space parameters – namely, smoothing intensity, meshing density, and partitioning size. Through a comprehensive analysis of over 1200 unique scale space constructions derived from a cohort of 185 aortic dissection patients, this work pinpoints optimal resolution scales at which shape variations are most strongly correlated with surgical outcomes. Importantly, these findings emphasize the pivotal role of a secondary discretization step, which consistently yield the most robust signal when scaled to approximately 1 cm. This approach enables the development of models that are not only clinically effective but also inherently resilient to biases introduced by patient population heterogeneity. By focusing on the appropriate intermediate scales for analysis, this study paves the way for more precise and reliable tools in medical imaging, ultimately contributing to improved patient outcomes in cardiovascular surgery.

## I. Introduction

THE clinical diagnosis and management of vascular diseases, particularly aortic disorders, heavily rely on the precise characterization of anatomical features. Current imaging-based diagnostic features predominantly focus on easily measured size metrics like maximum aortic diameter to provide a statistical measure to support surgical intervention. [1], [2], [3] Other metrics such as the tortuosity index aim to expand upon size information to include aortic shape, but these metrics have had little clinical impact due to their limited interpretations. [4] Unsurprisingly, for decades the most popular tool for identifying the complex geometrical nuances essential for risk assessment and treatment planning has been the trained eye of the medical professional. The eye has the innate ability to capture the wealth of geometric information from an image, simultaneously traversing the various scales within and allowing intuition to identify regions of interest. Even with the abundance of advanced methods which can provide detailed imaging with sub-millimeter resolutions [5], [6], modern computational tools are not fully utilized statistically, with most clinicians relying on their experience and intuition to make decisions. Likewise, despite technological strides in medical imaging and analysis, the heuristic nature of actionable signatures and parameter selections provided by these tools often limits their comprehension, widespread adoption, and more so the potential for robust and generalizable models driven by compounding statistical significance. Therefore, the most clinically useful anatomical features are those that most robustly quantify what the eye sees. This bridging from interactive and intuitive to analytical and statistical (qualitative to quantitative) is best hallmarked by the furthering development of automated image segmentation. [7], [8] Virtual anatomical models are becoming ubiquitous in clinical settings and complimentary computational methodologies are being developed to extract multiscale data from the model structures. These approaches are designed to not only replicate the multi-layered perception inherent to human observation but also provide the means to quantify the intrinsic geometric information within. Thus, by viewing aortic anatomy through this multiscale lens, we can gain detailed geometric insights that are otherwise hidden or obscured in single-scale observations, setting the stage for the introduction of computational tools designed to further explore and quantify the complexities of anatomical structures. [9], [10]

Recently, normalized fluctuation in integrated Gaussian curvature 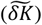 of 3D surface reconstructions of thoracic aortas using computed tomography angiography (CTA) imaging data was introduced as a size-invariant intrinsic shape quantity. 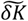 demonstrated enhanced classification in logistic regression machine learning (ML) models when fit to predict the success of thoracic endovascular aortic repair (TEVAR) for patients with aortic dissections (when coupled with aortic radius in a tandem size-shape feature space). [11] Briefly, the feature is calculated from CTA imaging by first approximating discrete point Gaussian curvatures (*k*_*g*_) for each vertex of a triangular 3D surface mesh using the discrete point curvature algorithm by Rusinkiewicz et al. [12] Then, the regions of constant Gaussian curvature are averaged together and used to calculate the total integrated Gaussian curvature (*K*). The fluctuation in *K* is *δK* = ⟨*K*^2^⟩ − ⟨*K*⟩^2^, thus describing the variation of integrated curvature values over the surface of the aorta, i.e., the “variation in bumpiness” or the magnitude of statistical deviation away from a perfect cylindrical aorta. While promising, the procedure we introduced to calculate 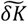 from CTA data involves three steps which all influence the scale space in which the feature is produced: *smoothing* of the aortic surface model (the removal of noise, but also structure) upon which a *triangular dense mesh* is applied (discretization of the surface) and subsequently *partitioned* (averaged) for local evaluations in integrated Gaussian curvature. In the original methodology, a heuristic approach was taken by implementing each of the three steps in a purely qualitative fashion, a position many clinician-scientists find themselves in: having access to an overwhelming number of advanced tools, but using mainly intuition to guide and/or optimize the generation of usable data. After all, the clinician traditionally uses their trained eye first to extract data from anatomical images. This is illustrated using the series of images in Fig. 1. Whether the surgeon sees the highly pixelated and rough unsmoothed segmentation surface of the aorta in Fig. 1.A or the integrated Gaussian curvature surface map in Fig. 1.E (or any of the images in between), their conclusions about the shape of the patient’s pathology and the impending implications for treatment remain the same.

**Fig. 1.**
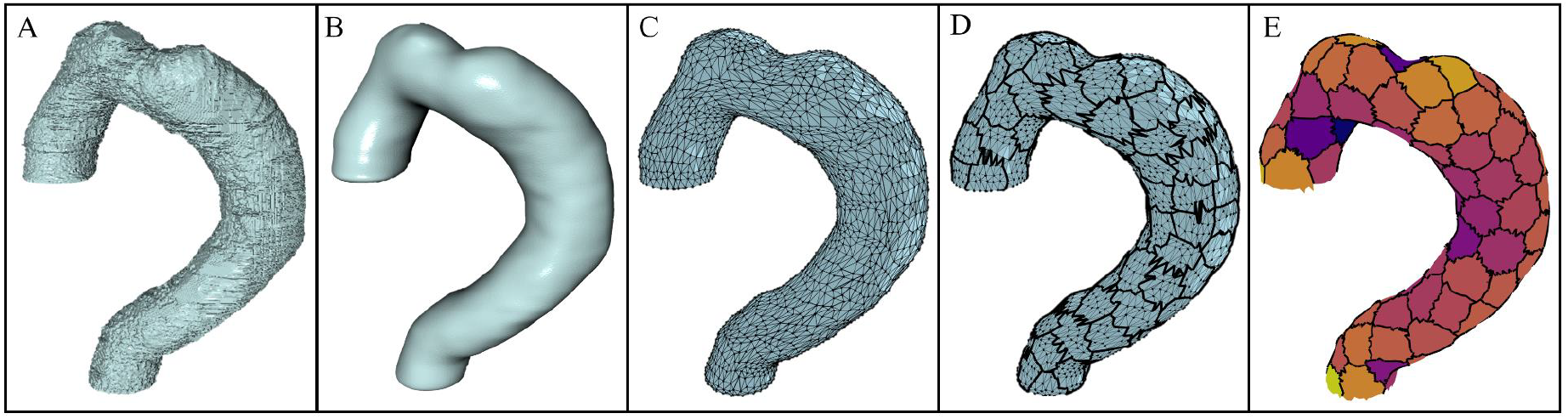
The evolution of a single aortic geometry through the subsequent steps in the methodology. (A) The raw segmentation mask produced directly from the DICOM image files with surface roughness from the image pixel resolution. (B) The smoothed segmentation mask after a recursive Gaussian smoothing operator was applied to the surface (the Gaussian length scale depicted in the image is 7 voxels). (C) The triangular surface mesh formed from the smoothed segmentation mask (5000 triangular elements are depicted). (D) The partitioned aortic surface after a K-means algorithm divides the mesh into a predetermined number of partitions distinguished by the dark borders (115 patches shown). (E) The total integrated Gaussian curvature (K) shown per-patch. The color of the patches now corresponds to the value of K, with lighter colors depicting regions of high K and darker colors depicting regions of lower K. Steps A-C are performed in Simpleware ScanIP and the steps D-E are performed in the 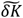-algorithm using Python libraries.

However, if a quantitative analysis route is taken, a plethora of challenges is introduced. Specifically, discretizing or taking derivatives on pixelated surfaces can introduce significant noise to calculations. Using the original paper as a prime example of an image-generated feature susceptible to these challenges, we employ here a data-driven methodology to identify the region in the scale space formed by smoothing, meshing, and partitioning which match clinical intuition and maximize the quantitative role of the feature in predictive algorithms.

This work is particularly influenced by the foundational theories of Witkin and Koederink on scale space, which describe the behavior of features as one varies the scale of observation. [13], [14], [15], [16] The primary goal of the scale space optimization is not only to enhance the signal-to-noise ratio but more importantly, to identify the scale, or rather the range of scales, in which the 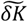 “event” occurs—the event being where the clinical utility is maximized, either for predictive algorithms or physical interpretation. For example, in the classic coastline measurement problem, it is well understood that the most effective measurements are not made by measuring grains of sand on the beach with a caliper or from taking low resolution pictures from the moon. Rather, the compromise between measurement length scale and resolution provides the best result. After all, the basic principles of cartography are strongly rooted in differential geometry and topology, which further link the instrumental works of Gauss to modern day scale space problems. By optimizing the scale space, smaller features like bays and inlets are smoothed away, coarsened over, or averaged out, while the general shape and length of the ‘coastline’ is maintained. Analogously, the operations of smoothing, coarsening, and averaging an aortic surface allow the removal of uninformative features from segmentations while capturing the variations in morphologic aortic shape.

The most commonly discussed operator throughout the scale space literature is the Gaussian function or diffusion equation. [17], [18] Diffusion only has the ability to destroy structure – any feature at a coarse level of resolution must also present at a finer level of resolution, while the reverse is not true, making the operator ideal for high-to-low resolution exploration.

Gaussian smoothing is also standard in data visualization and noise removal procedures, making it a suitable parameter for exploring the extent of segmentation annealing required to produce the best shape signal. Surface meshing is also ubiquitous in computational frameworks for modeling physical phenomena and can significantly simplify geometric calculations by discretizing complex surfaces into a collection of operable coordinates. [19], [20], [21] The literature on optimizing meshing for physical quantity estimation is rich but is simplified in this approach to provide the most generalizable results. In the following analysis, both extrinsic and intrinsic meshing discretizations are examined. However, while a fine-level of discretization is required to calculate *k*_*g*_ locally at a given mesh vertex, we showed previously that *k*_*g*_ alone does not produce a useful geometric signal in these data. Therefore, *k*_*g*_ is integrated over an additional length scale to produce a topologically invariant quantity which statistically describes the shape differences between TEVAR outcomes. The integration length scale, or the partitioning length scale, is an effective secondary coarsening and discretization of the initial mesh, now containing the average Gaussian curvature 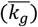 from a finer level of discretization and the partition area (*A*) at each partition centroid. Therefore, the per-partition integrated Gaussian curvature value is calculated as 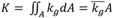 This highly effective down-sampling method averages out the noise from the per-vertex *k*_*g*_ calculation while capturing the shape over the surface area set by the partition length scale. This secondary coarsening strategy is often implemented when domain knowledge supports extracting features which exist from finer meshes at higher scales of measurement. [22]

Each of the three operators described serves a unique and pivotal purpose in quantifying shape variability on complex anatomical manifolds. Given there is not an a priori solution as to how shape variation should scale with aortic size or aortic disease, the signal produced must be characterized by its usefulness. The two most important characteristics of the 2D feature space are the size-shape invariance and the outcome class clustering, demonstrating the potential for high predictive accuracy and utilization in algorithms. Using metrics which score these attributes, a vast combination of datasets constructed using different Gaussian smoothing operator length scales (noise removal), meshing density (fine-discretization), and partition size (coarse-discretization) are evaluated. More importantly, the ranges over which a high quality 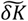 signal appears is designated a “stable zone” for feature extraction (with intentional reference back to Witkin’s idea of “stable features”). [14] Furthermore, the stable range of length scales in turn produce a stable range of viable solutions, with each aorta having a unique shape value for each stable parameter combination. This effective up-sampling of the data over all useful scales of observation may in practice mimic the multilayered perception of the human eye and in turn, may provide computational methodologies with data that best represent what the trained professional’s eyes really see. The originating work introducing 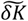 as a descriptor succumbed to a heuristic methodology to convert the qualitative but rich signal into a useful quantitative measure, not being carefully tuned for proper data-driven usage. Indeed, while achieving 92% classification accuracy on preoperative *and* postoperative data, the accuracy decreased to just 61% when just preoperative imaging of the patient anatomy was used. Aside from leveraging postoperative data when evaluating the 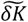 signal throughout scale space, only preoperative data is used within the classification models to properly demonstrate the predictive utility of the feature in an environment with true clinical translatability.

## II. Methodology

### A. Imaging Data and Segmentation

A data cohort of 185 unique patients and 380 corresponding CTA scans were collected for analysis from the Human Imaging Research Office (HIRO) at the University of Chicago. All data collection and analysis were performed following the guidelines established by the Declaration of Helsinki and under institutional review board approval (IRB20-0653, IRB21-0299). Each patient and their corresponding scans are labeled using one of three discrete levels: nonpathological, successful TEVAR, or unsuccessful TEVAR (surgical reintervention or death one-month post operation), which is analogous to the labeling scheme in the original analysis. Data were received in the form of deidentified digital imaging and communications in medicine (DICOM) files from CT instrumentation spanning a variety of models and resolutions; the range of xy-spacing was 0.3-0.8 mm and z-spacing was 0.3-1.0 mm. Only 3D scans were used in building the dataset and all chest CT scans were obtained from the axial cut of the original DICOM file. Surface models of each unique anatomy were created from the files using Simpleware (S-2021.06-SP1, Synopsys, Mountain View, CA) software. Segmentation, the reconstruction of the 3D thoracic aorta, was performed semi-automatically. A 3D mask that best captured the aortic anatomy was generated by thresholding and adjusting the upper and lower bounds of the greyscale DICOM series, capturing both the aorta and residual tissue for each slice. Then, slice by slice, the outer perimeter of the aorta was identified, removing noise, residual tissue, and branch vessels. The proximal and distal boundaries of the mask were defined by the aortic sinus and celiac trunk, respectively, and any aorta beyond those bounds was completely removed from the mask. The segmenter would then alternate between the visualized 3D model of the segmented aorta with the axial cut, sagittal cut, and coronal cut of the CT scan and continue to refine the raw segmentation to best capture the aortic anatomy. Lastly, removal masks would be generated at the proximal and distal bounds to precisely define the ends of the 3D model (Fig. 1.A).

### B. Smoothing, Meshing, and Partitioning

Smoothing of the raw segmentation mask is a necessary step to eliminate the pixelated surface structure generated from the image processing (Fig. 1.B). The smoothing process employed was comprised of two independent steps: mask dilation and recursive Gaussian filters. To study the implications of continuously broadening Gaussian smoothing, the length scale of the 3D isotropic Gaussian surface function applied was varied from one to ten voxels in intervals of one (*σ*_*x*_, *σ*_*y*_, *σ*_*z*_ ∈ {1, 2, 3, … 10}). Constant dilation was implemented equally across all aortas to ensure the smallest aortas (with the least voxels) maintained their original topology during the smoothing process. The dilation filter applied grew the segmentation masks by adding 2 outer layers of voxels to the surfaces, which provided sufficient additional mass to preserve the smallest aorta’s topologies during the most intense smoothing.

As each of the 380 segmentation masks was smoothed by varying degrees, each was subsequently meshed using 5 triangular mesh densities (1, 5, 10, 50, 100 *mm*^2^) using Simpleware’s default meshing algorithm. Each mesh element density was kept approximately constant by constraining the Simpleware meshing algorithm to a narrow range of optimization parameters, however some minor modifications to a pure lab-frame extrinsic mesh were enforced by the software along sharp edges or creases. Subsequently, each 1 *mm*^2^ element mesh was decimated (using open-source Python libraries for meshing, TriMesh and Open3d) to an intrinsic mesh with a constant number of elements. [23], [24] The quadric error metric decimation algorithm was used to create a variety of constant element meshes which included 50, 100, 500, 1K, 5K, 10K, 50K elements (Fig. 1.C). The algorithm is designed to prioritize keeping distinct shapes and features over subsequent simplifications of the mesh, to perform the optimized coarsening. [25] Visual differentiation of the extrinsic and intrinsic meshes are shown in Fig. 2.

**Fig. 2.**
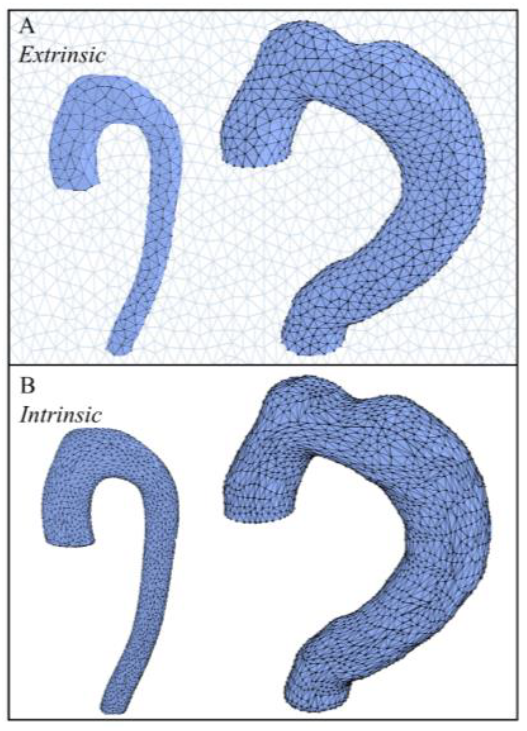
Images extracted for both (A) extrinsic and (B) intrinsic meshes. The extrinsic meshes are created within the Synopsis Simpleware software and are set to a variety of mesh densities (1, 5, 10, 50, 100 *mm*^2^ elements). In an approximate lab-frame approach to meshing, the number of elements on the surface of the manifold scales directly with the surface area of the anatomy itself, therefore providing a curvature evaluation (*k*_*g*_) over a fixed physical length scale, set by the density of the mesh. The intrinsic meshes are created by decimating the highest density extrinsic mesh using the Trimesh package in Python. Each is targeted to have a constant number of elements regardless of the size of the anatomy and span (50, 100, 500, 1K, 5K, 10K, 50K elements). Within these meshes, *k*_*g*_ is calculated over a relative distance which increases as the number of elements decreases.

Each of the uniquely smoothed and meshed datasets were then run through the discrete point curvature algorithm to calculate per-vertex Gaussian curvatures (*k*_*g*_) from the two per-vertex principal curvatures (*k*_1_, *k*_2_). [12] Subsequently, the 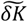-algorithm previously mentioned was used to calculate the shape metric and the mean root Casorati curvature ⟨*C*^1/2^⟩ where 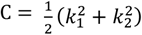, which is an analog of mean inverse aortic radius (the size feature in the 2D size-shape feature space). The final parameter in the analysis, the partitioning length scale, was varied within the 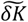-algorithm for each dataset. Each mesh, at this point with per-vertex Gaussian curvatures, was divided into either a fixed number of surface partitions *p* = 10, 50, 100, 150, 250 or the number was calculated using the scaling relationship 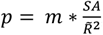, where *SA* is mesh surface area and 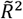 is the median radius squared, to achieve floating partition totals that scale with aortic size (Fig. 1.D). In the original analysis the prefactor (*m*) was set to 1, but here it is varied from 0.1, 0.5, 1, 5, and 10. Finally, for each aorta, in each scale space dataset, a 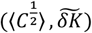 feature pair was calculated.

### C. Scale Space Evaluation

Each dataset was then evaluated using three different characteristics of the resulting 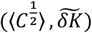 feature spaces. First, each dataset was fit with a generalized power function 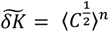 (i.e., *shape* = *size*^*n*^) where *n* ∈ ℝ and such that *n* = arg min 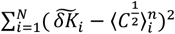. Each fit produced a coefficient of determination (*r*^2^) value which was used as the scoring metric. Second, a classification algorithm was fit using multinomial logistic regression and was used to predict the outcome labels as a function of the 2D size-shape feature space. The learned decision boundaries determine per-point classification and the F1-score for the data cohort was calculated. The F1-score is the harmonic mean of the precision and recall of the prediction accuracy and is used to quantify how linearly separable the clusterings of outcomes are while also considering class label imbalances. Lastly, a K-means algorithm was used to separate each dataset into *k* = 3 clusters which is the ground-truth number of discrete labels within the dataset. However, the model assigns each 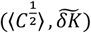 pair of datapoints to an arbitrary group 1, 2, or 3 using only the spread of the collection amongst the size-shape space. The agreement between the clustering model and the ground truth labels is then evaluated using the Adjusted Rand Index (*ARI*). [26] The Rand Index (*RI*) is calculated by taking the fraction of all pairs of samples that are either in the same cluster in both clinical and model labels or in different clusters in both clinical and model labels. The *ARI* is adjusted for chance and is computed as

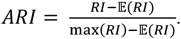

After each metric (*r*^2^, *F*1, *ARI*) was calculated, they were each scaled from zero to one and the values were added together to form a standardized and lumped cumulative score for evaluation. The two clustering metrics were halved as to not bias the cumulative score towards clustering over power law fitting, while maintaining independent representation of linear-separability (logistic regression) versus nonlinear-aggregation (K-means clustering). The cumulative score is thus 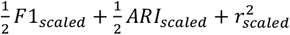 and is bound between zero and two. Once a cumulative score has been calculated for each dataset, a virtual grid is constructed which spans broad ranges along the Gaussian smoothing operator length scales, meshing density (fine-discretization), and partition size (coarse-discretization). A total of 1200 differently scaled datasets were achieved by spanning 10 smoothing length scales, 12 meshing discretizations (5 extrinsic and 7 intrinsic), and 10 partitioning discretizations (5 static and 5 floating). A schematic overview of this scale space grid methodology is detailed in Fig. 3.

**Fig. 3.**
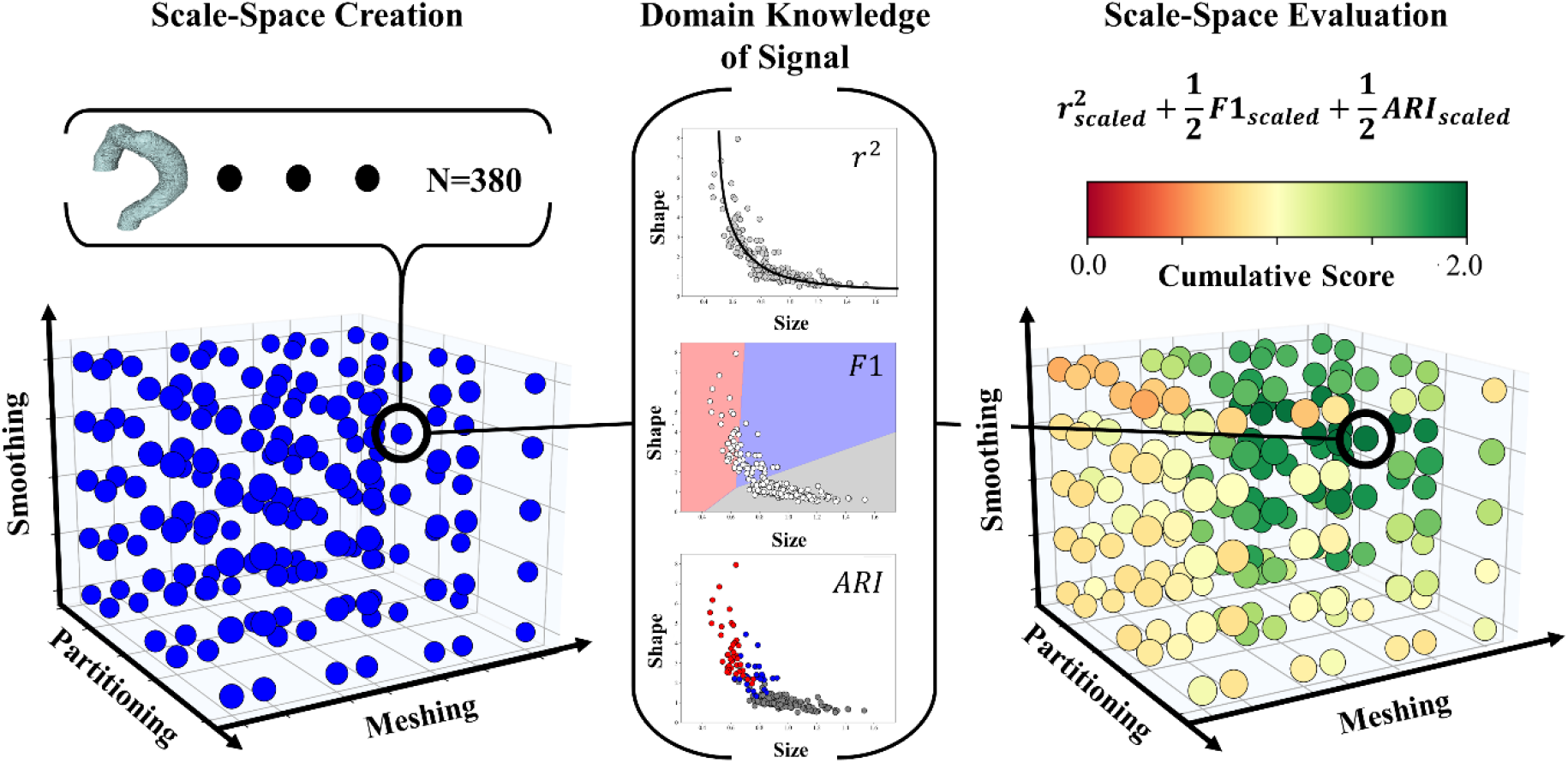
Schematic overview of the methodology for evaluating the scale space for aortic anatomy. During creation each of the 380 segmentation surfaces is subsequently smoothed, meshed, and partitioned using a variety of length scales along each of the 3 axes. Each discrete blue datapoint in the left-hand figure is itself a dataset containing the 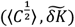 feature pair for each of the 380 aortas from just one of the four primary/secondary discretization combinations. The image represents 133,000 aortic scans in total (10 smoothing variations, 7 meshing variations, and 5 partitioning variations). Overall, 456,000 aortic scans were evaluated amongst the four scale space cubes created (133,00 from each of the two *I* methods, and 95,000 from each of the two *E* methods). Depicted in the image is just one of these four cubes. For each of the datasets within the cube, the signal evaluation process in which clinical domain knowledge is used to estimate the usefulness of the signal produced. A power-law r^2^ value, a classification F1-score, and a clustering ARI score are all calculated from the 2D scatter data. In the final step, the scale space is evaluated by taking the cumulative lump score of the three metrics and reassigns the grid position with the score. The gradient in scores across the grid can therefore be visualized and provide an illustration to how the stability of the signal changes in all 3 dimensions formed by the scale imposed during dataset creation.

In addition to evaluating the scale space using the cumulative scores, the improved utilization of the tuned feature space is further assessed through the application of regularized multinomial logistic regression and Gaussian process classification (GPC). [27] These two widely used predictive ML models offer complementary perspectives on the data, with logistic regression providing a linear approach and GPC offering a more flexible, nonlinear methodology. The logistic regression model was fit in a manner consistent with the procedures described in the original work, involving careful preprocessing, validation, and regularization steps to ensure robustness and prevent overfitting. This model serves as a baseline for evaluating the linear separability of the classes within the optimized feature space. The GPC model, on the other hand, leverages the power of Gaussian processes to model complex, nonlinear decision boundaries. Specifically, the GPC model was fit using an anisotropic radial basis function (RBF) kernel, which allows the model to adapt to the specific geometry of the data. The RBF kernel is particularly effective in capturing local variations within the feature space, making it well-suited for datasets where class boundaries are not linearly separable. The anisotropic nature of the kernel means that it can adjust its shape according to the distribution of the data, providing a tailored fit that reflects the underlying structure of the feature space. To visualize the model’s performance, per-class probability contours were generated and constrained to regions within one standard deviation of the available data points. This constraint ensures that the visualized decision boundaries are meaningful and closely aligned with the actual data, avoiding extrapolation into regions where the model has insufficient information. These probability contours offer a visual representation of the regions where the GPC model predicts each class with high confidence. One of the key advantages of GPC is its ability to quantify uncertainty in label assignment. This is particularly valuable in medical imaging analysis, where understanding the reliability of predictions is crucial. The GPC model inherently provides a measure of uncertainty by generating probabilistic outputs rather than hard classifications. This allows for the identification of regions in the feature space where the model’s predictions are less certain, often corresponding to areas where the classes overlap or where data density is low. These regions of uncertainty are directly compared to the stability regions identified during optimization, providing a comprehensive view of the model’s reliability and the robustness of the feature space.

## III. Results and Discussion

As described in the methodology, 1200 unique scale space constructions of the TEVAR dataset were evaluated during the optimization. Within those datasets, four unique parent methodologies can be categorized: extrinisc (E) or intrinsic (I) meshing, and fixed (F) or scaling (S) partitioning. For the remainder of the discussion these four groups will be referred to as E-F, I-F, E-S, I-S. Table I provides a comprehensive list of outcome metrics for each of the best performing parameter sets for the 4 parent methods.

**Table 1.** Scale Space Optimization Results.

| Method | Smoothing | Meshing | Partitioning | $r^2$ | F1 | ARI | Cum. Score | Partition Area [mm<sup>2</sup>] | Elements per Partition |
| --- | --- | --- | --- | --- | --- | --- | --- | --- | --- |
| **E-F** | 7 vox. | 1 mm<sup>2</sup> | 250 | 0.57 | 0.86 | 0.74 | 1.65 | 137 ± 75 | 322 ± 182 |
| **E-S** | 7 vox. | 10 mm<sup>2</sup> | $1 \cdot \frac{SA}{R^2}$ | 0.76 | 0.92 | 0.79 | 1.91 | 252 ± 169 | 58 ± 39 |
| **I-F** | 7 vox. | 5000 e | 250 | 0.60 | 0.88 | 0.82 | 1.80 | 137 ± 79 | 19 ± 1 |
| **I-S** | 7 vox. | 5000 e | $1 \cdot \frac{SA}{R^2}$ | 0.80 | 0.91 | 0.83 | 1.89 | 248 ± 166 | 32 ± 5 |

Summary metrics for the optimal case from each of the 4 unique parent methods. For each method the optimal Smoothing, Meshing, and Partitioning value is reported alongside the corresponding $r^2$ , ARI, and F1 score values. Smoothing is reported as the recursive Gaussian operator length scale which is the width of the Gaussian kernel applied to the surface. In the z-direction of the scan the average z-spacing was 0.6 mm (e.g., a smoothing length scale of 7 converts to 4.2 mm on average). The average ± standard deviation for the partition surface area and the mesh elements per partition are also reported. The cumulative scores of each method are scaled from 0 to 2.

The results in Table I highlight several key findings from the methodology analysis. The optimal length scale for the recursive Gaussian smoothing filter was consistently identified as 7 voxels (average length of 4.2 mm). Additionally, in all four scale space grids (Fig. 4), the most stable regions of the grid are all centered about the 7-voxel smoothing length scale suggesting that at this level of surface diffusion lower level structures that have been decimated are not informative in predicting aortic disease state. Similarly, at smoothing length scales greater than 7 voxels, important shape information as it pertains to aortic disease state begins to be destroyed. Of particular note are the similarities between the physical thickness (intima, media, and adventitia) ~*O*(1 *mm*) and the optimal smoothing length scale. [28] The similarity suggests that aortic surface structures sensitive to thickness likely do not play a role in characterizing global aortic shape.

To identify the optimal method in each category, the parameter groups that achieved the highest cumulative scores were first selected and among these, the simplest mesh was chosen. For instance, within the E-S group, both the 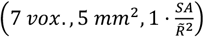 and the 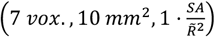 configurations achieved top scores of approximately 1.91. The latter was selected because it is a coarser mesh. The preference of the simpler mesh is a minor and more practical point because if a range of scales produce the desired signal, and only one is to be chosen from the range, it only makes sense to choose the most computationally efficient one.

Fig. 4 visualizes this methodological scale space grid for the four parameter sets (I-S, E-S, I-F, and E-F) used in the optimization process. The figure demonstrates that high-quality signals were obtained over the ranges 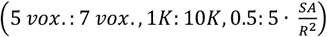 for I-S (Fig. 4.A) and 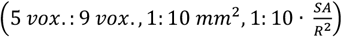 for E-S (Fig. 4.B) while the I-F (Fig. 4.C) and E-F (Fig. 4.D) approaches had less than optimal results. A significant finding is the notable difference in power law fitting between methods employing a scaling number of surface partitions. Fig. 5 presents the logistic regression outputs for the preoperative-only datasets for each optimal method. A key observation from the high-quality signals in both Fig. 5.A and Fig. 5.B is the anisotropic distribution of nonpathological aortas, which aligns with the expectation of shape-preserving growth during healthy tissue development. The shape of the 2D signal resembles an “elbow,” where the introduction of disease results in an upward trajectory on the shape axis. This characteristic is crucial, as capturing the absence of shape variability is just as important as detecting its presence. The E-S (Fig. 4.B and Fig. 5.B) method stands out amongst the rest for three reasons: 1) the means of the three groups are the furthest apart – nonpathological 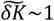, successful TEVAR 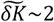,unsuccessful TEVAR 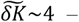 along the shape axis; 2) nonpathological aortas exhibit minimal shape variation; and 3) the classification score is the highest among the methods, achieving on average 71% and 88% accuracies for the successful and unsuccessful TEVAR cases, respectively. In direct comparison to the original work and model using 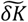, this improved scaling imposed during the calculation results in an 83% accuracy when tasked with predicting TEVAR outcome, compared to 61% previously. Given the optimal primary and secondary discretizations from the E-S method, the inner scale imposed by the mesh was found to be 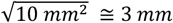 while the outer scale imposed by the partitions was 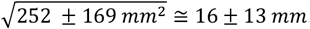.These scales correspond well to the physical characteristics of the aorta, which has a thickness ~*O*(1 *mm*) and a radius ~*O*(10 *mm*). [28], [29] This alignment only further emphasizes the importance of anatomical length scales when extracting geometric information from segmentation surfaces.

**Fig. 5.**
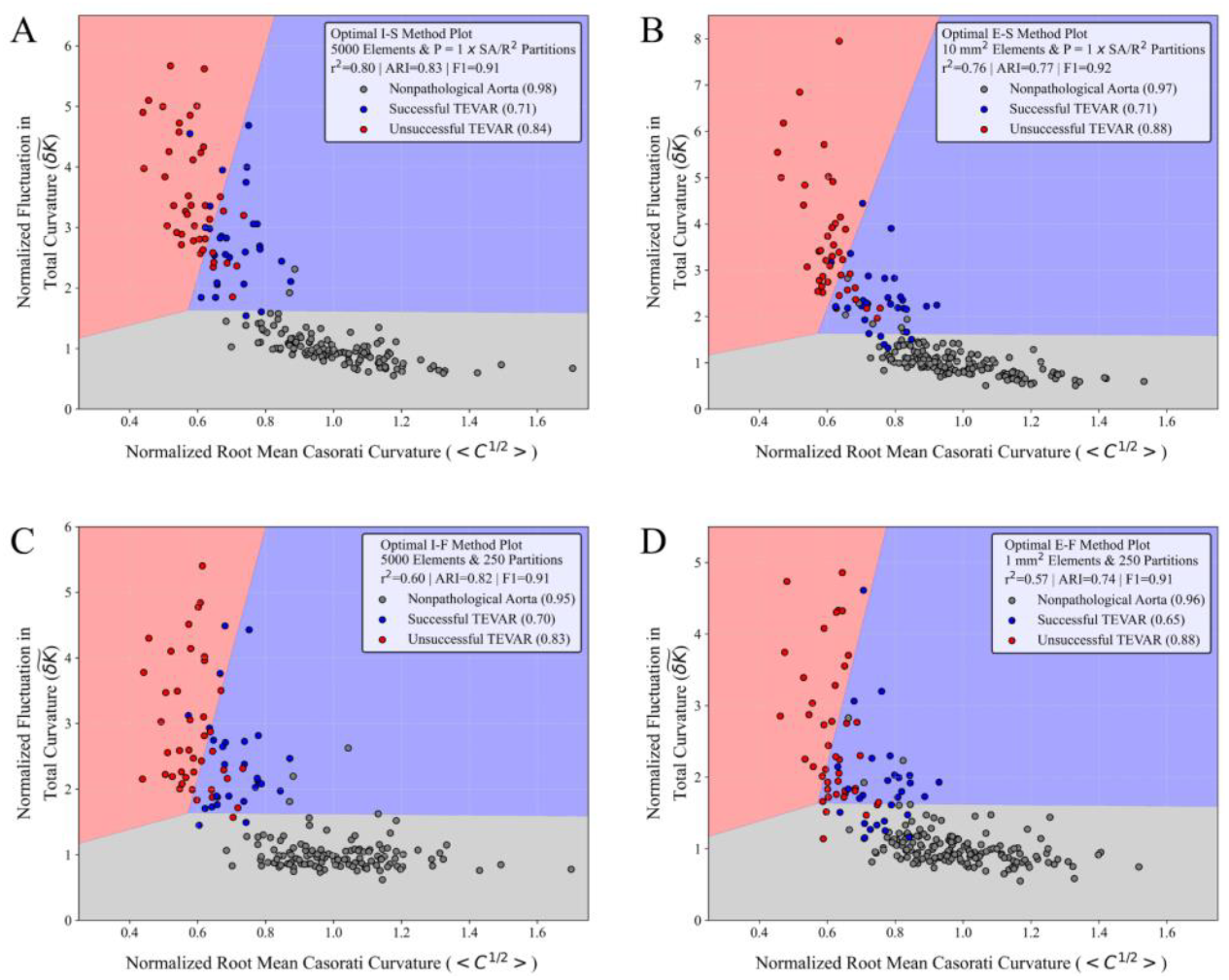
The optimal size-shape feature space and corresponding multinomial logistic regression decision boundaries for the (A) I-S, (B) E-S, (C) I-F, and (D) E-F approaches. Both axes in all panels are normalized to the mean value of all nonpathological points in each dataset respectively (i.e., the point [1, 1] depicts the average normal thoracic aorta in both size and shape). The legend in each panel describes the method used to produce the optimized result, the mesh and partitioning scales used, the value of each of the three in-part scoring metrics, and the per-class percent accuracies from the linear decision boundaries averaged over 50 unique train-test splits of the dataset before model fitting and regularization.

To further evaluate the robustness of the E-S methodology using only preoperative data, the feature pair average from each unique scale combination which obtained a cumulative score > 1.85 set was computed. Fig. 6.A shows the averaged points with error bars representing the standard deviations in both dimensions. These error bars quantify the variability inherent in the “stable” zone of the data, indicating the extent to which variations in smoothing, meshing, and/or partitioning parameters influence the size and shape metrics while still providing a high-quality signal. Overlaid on this scatter plot are class contours obtained from a GPC model. These contours delineate the nonlinear decision boundaries between different classes (e.g., nonpathological, successful TEVAR, and unsuccessful TEVAR). It is noteworthy that the GPC contours closely align with the variance error bars, demonstrating the model’s capacity to capture not only the central tendencies of the data points but also their potential variability due to slight parameter adjustments. This alignment suggests that the GPC model is well-suited for accommodating the inherent uncertainties in the data, offering a probabilistic framework that effectively accounts for small-scale variations. An additional observation from Fig. 6.A is the distinctive pattern of error bars across the different aortic categories. For nonpathological aortas, the error bars are large in the size dimension but small in the shape dimension, reflecting stable shape characteristics despite variations in size. Conversely, for preoperative CTA scans that would later become unsuccessful TEVAR cases, the error bars are large in the shape dimension and small in the size dimension, indicating significant shape variability. This pattern suggests that as disease progresses, the variability in shape becomes more pronounced and presents at a wider range of length scales, which aligns with the understanding of disease progression as increasingly non-uniform and potentially chaotic. [30], [31] The isotropic error bars observed in eventually successful TEVAR cases further emphasize the balance between size and shape variability, perhaps indicative of a more controlled clinical outcome where aortic morphologies reach anatomic stable points even in the setting of disease. [32], [33] The pattern is also duplicated, if not further developed, when the horizontal axis is converted back to aortic radius from inverse size in Fig. 6.C. The original choice to use inverse size was done to emphasize the invariance and asymptotic nature of the 2D inverse size-shape signal, and it certainly served a role in this analysis. However, it may be more interpretable to visualize the results of future scans in the noninverted space. Furthermore, it may enhance performance of predictive algorithms as it provides more space for the pathologic groups to occupy along the size axis, demonstrated by the 6% and 10% successful TEVAR class accuracy increases between Fig. 6.A&C and Fig. 6.B&D, respectively.

**Fig. 6.**
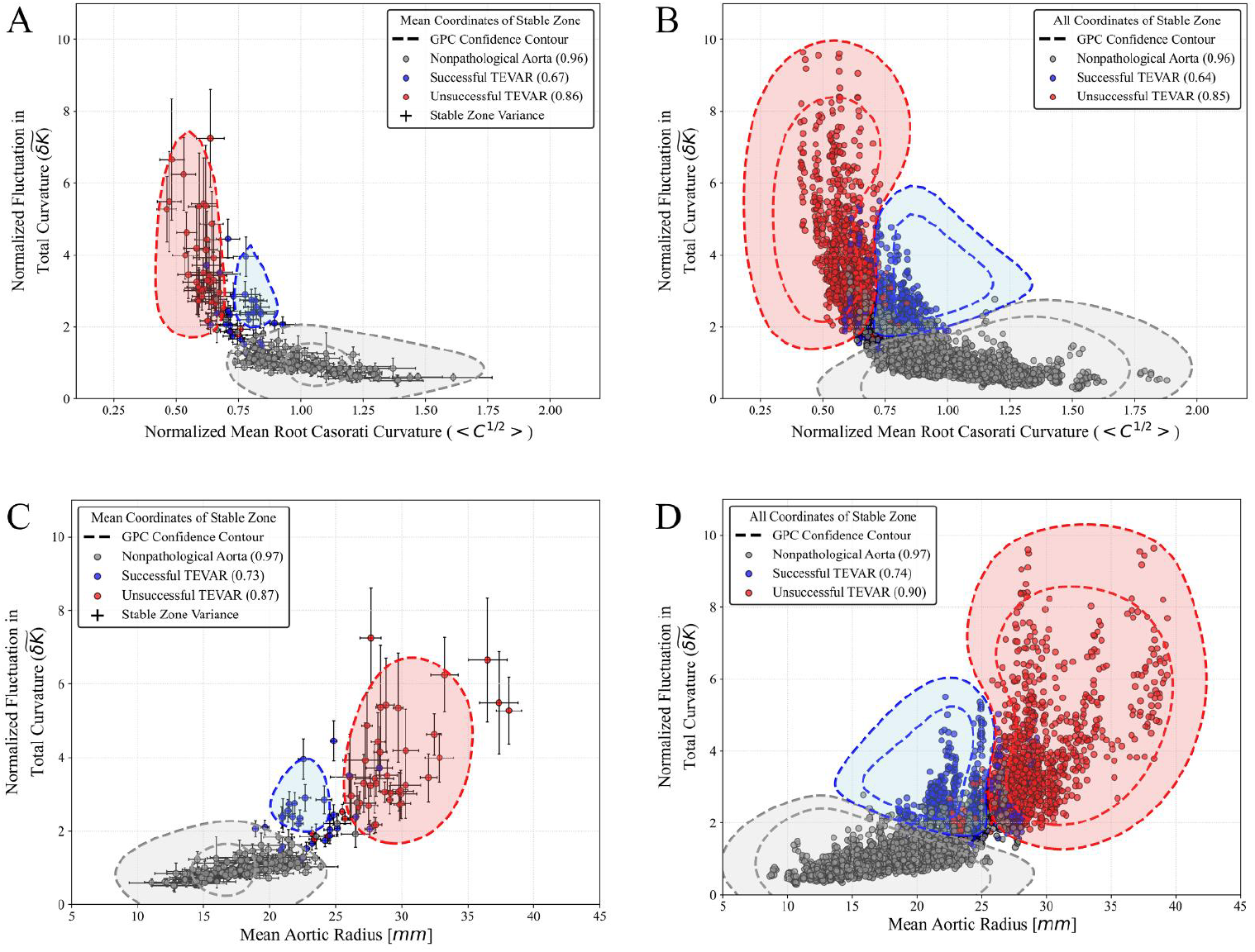
Gaussian process classification (GPC) classification boundary plots for the dataset formed from the means of all E-S method outputs with cumulative scores greater than 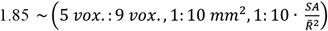.Each panel depicts at least a 50% (outer) and for some others an 80% (inner) confidence contour. (A) Each data point error bar depicts the standard deviation in both the inverse size 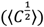 and shape (*δK*) axis respectively for the collection of “stable” instances of that scan that formed the mean. (B) The same analysis as in (A) however, with each instance from the stable collection being represented as its own discrete point. (C) The panel contains the exact same data as in (A) but with the x-axis transformed into mean aortic radius in units of millimeters. Similarly, in panel (D) the data replicates what was done in (B) but in the radius-space. In plots (A) and (B), both axes are normalized to the mean value of all nonpathological points in each dataset respectively while in plots (C) and (D) normalization is not performed on the x-axis to keep the magnitude of the size values directly interpretable. The legend in each panel contains the per-class percent accuracies from either the nonlinear (GPC) decision boundaries.

In Fig. 6.B, the analysis is expanded to encompass all data points within the >1.85 cumulative score stable zone contributing to the biaxial variance discussed above. Instead of relying solely on mean points, the GPC model is fit on all stable zone points, reflecting variations in smoothing, meshing, and partitioning parameters over the range of corresponding length scales. This represents a novel scale space-based up-sampling method not explored in the cardiovascular imaging literature. The resulting GPC contours, while similar in location and shape to those in 6.A, are more expansive, indicating that the incorporation of additional data points leads to broader, more robust decision boundaries most notably demonstrated by the occurrence of 80% confidence regions (the inner of the two ovular contours depicted for each class label). The broader contours indicate that the GPC model, when trained on a more comprehensive dataset, capture the full range of signal length scales, while only fluctuating marginally in prediction accuracy. Each data point in this expanded and up-sampled dataset represents a slightly different scale of extraction of the original data, thus reflecting the multiscale nature of the observed phenomena. Given the principles of scale space theory, the figure underscores the potential of such a methodology for up-sampling (or perhaps better coined ‘scale’-sampling). By considering multiple length scales of the same data, the GPC model becomes more robust, capturing subtle variations that may be overlooked when working within a single scale. Fig. 6.D shows the scale-sampled dataset in radius-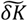 space and demonstrates perhaps the most useful and translatable output from this analysis, obtaining the best accuracy scores and most robust decision boundaries, while also being clinically actionable.

Fig. 7 presents the same data as Fig. 6 but uses logistic regression instead of GPC. Despite the regression model being a simpler linear classifier, it produces classification contours that are consistent with those generated by GPC, specifically and most importantly at the class interfaces. This result is important for two reasons. First, it demonstrates that the linear model can adequately (within a few percentage points in accuracy) capture the essential structure of the data, even in the presence of scale-induced variability. Second, it suggests that while the GPC offers a more nuanced, probabilistic approach, the overall classification outcome is sufficiently robust that simpler models can achieve comparable results. Lastly, the potential for scale-sampling through multiscale data points presents a promising avenue for enhancing model performance, particularly in datasets where capturing subtle differences is crucial. As shown by comparing Fig. 7.A&B to Fig. 7.C&D where Gaussian up-sampling is performed, the multiscale approach provides a significantly more representative result of a larger population of data than a purely statistical approach to up-sampling.

**Fig. 7.**
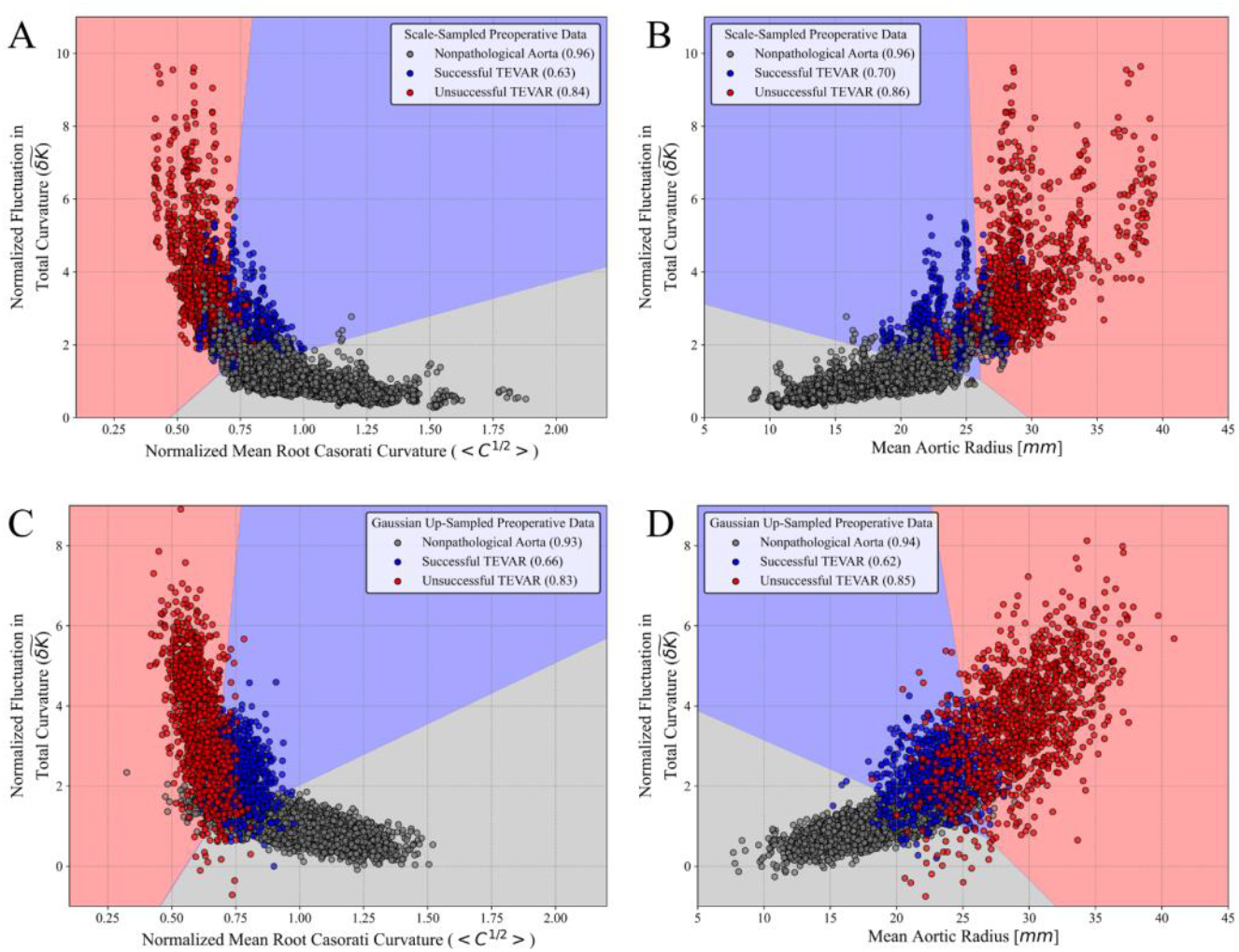
Logistic regression boundary plots for scale-sampled (A and B) and Gaussian up-sampled (C and D) datasets. The data points in (A) and (B) are identical to those shown in Figure 6.B and 6.D, respectively. However, the decision boundaries are now modeled linearly using regularized logistic regression. Each set of decision boundaries shown is the optimal output from 50 random initializations of training and testing data when fitting the regression. For comparison, the data points in (C) and (D) are up-sampled using a Gaussian distribution probability density function. For each class label, the bivariate means and covariances were calculated from the data in Figure 6.A and 6.C respectively and used to fit a Gaussian. Each PDF was subsequently up-sampled proportionately to match the number of per-class data points in (A) and (C) for direct statistical and spatial comparison between the two sampling methods. Analogously to Fig. 6, *δK* and 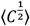 are normalized to the mean nonpathological scan values, while radius is left unnormalized for direct interpretation.

## IV. Conclusion

The scale space analysis conducted on the normalized fluctuation in integrated Gaussian curvature 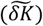 feature significantly enhanced signal strength and predictive accuracy in classifying aortic dissection intervention outcomes. Notably, the predictive performance using only preoperative data far surpasses that of our previous work (83% versus 61% in TEVAR outcome prediction). The ability to derive such a strong signal using only preoperative imaging transforms this model into a translatable decision-making algorithm. By shifting away from heuristic methodologies that have traditionally dominated the field, this work demonstrates the strengths of employing scale space analysis on imaging-sourced data. Specifically, the findings suggest that engineers and scientists developing medical imaging analysis tools possess considerable flexibility in data preprocessing. Whether it be through structural simplification via smoothing, discretization through meshing, or scale imposition through partitioning, the value of investigating the roles of resolution and scale in imaging data is invaluable. These findings also suggest, rather intuitively, that the easily measurable characteristic length scales of the system are a good place to start. In investigating the length scale of disease signature in patients with thoracic aortic dissections, we found that smoothing and discretization on the order of anatomical thickness along with signal measurement on the order of anatomical radius provided the most robust preoperatively accessible signal. The partitioning step sets the *inner scale* of the signal and provides access to a feature of *normalized shape density*. The simplicity of the optimal approach, coupled with its effectiveness, highlights the potential for these methods to be broadly adopted in clinical settings without sacrificing accuracy or interpretability.

In the broader context of medical imaging, practitioners often have limited control over the outer and inner scales of the image—the window set by the clinician and the resolution determined by the imaging tool, respectively. In contrast, the human eye collects data by operating on a variety of scales while simultaneously filtering out uninteresting structures. Therefore, the challenge in modern medical imaging analysis lies in identifying the right scale(s) for performing quantitative methods. This study contributes to that goal by demonstrating the importance of scale space evaluation and by providing a framework for its optimization through data-driven methods.

Finally, it is important to recognize that without prior knowledge of specific features of interest, it is challenging to carry out the methodology detailed above. However, by integrating domain knowledge from biology, anatomy, and perhaps most importantly clinical intuition, we can carefully guide computer-driven methods into quantifying the difference between two objects that “look” different. The combination of this knowledge with statistical reinforcement of shape information that successfully distinguishes different anatomies offers a powerful guide for optimizing the scale at which data analysis should occur.

## Data Availability

All data produced in the present study are available upon reasonable request to the authors. The imaging data reference in the manuscript is available at https://github.com/SurgBioMech/plos_data

